# Methods and Outcomes Reporting in Exercise-Based Cardiovascular Rehabilitation Research: a cross-sectional analysis

**DOI:** 10.1101/2022.07.21.22277905

**Authors:** Guilherme W Freccia, Rafaella Zulianello dos Santos, Leonardo De Lucca, Andrea Schaefer Korbes, Tales de Carvalho

## Abstract

The poor reporting quality of methods and outcomes is relatively recognized in the biomedical field. Its prevalence and implications have been studied in the cardiovascular rehabilitation literature but not so extensively in exercise-based trials. Our main objective was to cross-sectionally estimate the prevalence of both methodological and outcome reporting items in CVR trials with EBI. We also searched for associations (secondary outcomes) between (1) the effect size reported and the direction of the primary outcome, as well as (2) associations with the frequency of Spin. We cross-sectionally screened the sample of eligible trials dated between 2017 and 2021, and then collected the prevalence of methodological and outcome characteristics, independent and blinded manner. Our study shows that there was an insufficient reporting of methods and outcomes. Also, studies reporting effect size measures had a lower chance of Spin. The primary outcome effect size was not reported in 35% of the studies SES. However, more than 2/3 of the sample (69%) had a statement in the discussion or conclusion sections mentioning clinical relevance or meaningful benefit of the statistically significant results. Selective outcome reporting has important implications for translating science into practice, once not so threatens the validity of an intervention effectiveness, but also frustrates the use of its evidence in meta-analyses.

**What is new?:**

1. Our study shows that randomized controlled trials with cardiovascular rehabilitation based on exercise insufficiently reported the various methods and outcomes characteristics.
2. Although nearly 70% studies had stated its outcomes as clinically meaningful within our sample, about 41%of the studies clearly stated the primary outcome confidence intervals.
3. More than half of the sample presented at least one spin in the results section, and studies reporting effect size measures had a lower chance of Spin.
4. 40% of the studies within our sample did not report *a priori* sample size calculation, with 1/4 not stating the number of randomized subjects that could meet the intended power.
5. We did not find any associations regarding the direction of the results (positive or negative) and the prevalence of spin, contrary to what have been found in the literature.

## Introduction

Publishing peer-reviewed scientific reports in biomedical journals is still the most recognized pathway to disclose health scientific research. Research quality (i.e., how detailed and transparent the information about the design, conduction, and analysis of a study is) supports replication and helps to ensure internal and external validity. [1,2]. However, even when good quality reporting takes place, methodological misconduct could mask the credibility of the findings [3]. As an example, the absence of allocation concealment might bring high risk of selection bias to randomized controlled trials (RCTs), which can be associated with inflated estimates of outcomes in favor of the experimental group [4]. In order to assess the quality of a scientific report, we should carefully take into consideration not just the study design and conduction (methodological quality), but also the proper communication of the results to avoid selective outcome reporting [5,6].

The poor reporting quality of methods and outcomes is relatively recognized in the biomedical field. Despite the availability of checklists and reporting guidelines [7–9], cornerstone items for the internal validity of a RCT have been poorly reported [8,10,11]. At the same time, we do not know whether the authors really conducted a specific method, or if it was simply not reported, for whatever reason. In the field of cardiology, randomization, blinding, allocation concealment and sample size are some of the most compromised methodological items in terms of reporting [12–14]. More specifically, studies of cardiovascular rehabilitation (CVR) based on exercise had its risk-of-bias assessed in two Cochrane Collaboration reviews [15,16], which presented some concerns regarding biases such as: selection (allocation and randomization), detection (blinding of outcomes), and attrition (incomplete outcome data).

Statistical reporting issues have gained attention as the worrying about the prevalence of underpowered RCTs increase. For example, inadequate description of statistical power calculations [7], inappropriate choice of dispersion and precision measures [17], and the significance level adopted [18–20], may have affected the overall quality of systematic reviews with meta-analysis (SRMA) that we rely when clinical decisions need to be taken [21]. The prevalence and implications of such methodological features have being studied in the CVR literature [22–24], but not so extensively within exercise-based intervention (EBI) RCTs, which supports the objective and primary outcome of the present study: to cross-sectionally estimate the prevalence of both methodological and outcome reporting items in CVR trials with EBI. For our secondary outcome, we searched for associations between (a) effect size reporting prevalence and (b) the reported primary outcome direction (i.e., hypothesis confirmation or refutation) along with the prevalence of Spin.

## Methods

This is a cross-sectional study design with a systematic sample of CVR trials containing EBI, which had its reporting of both methods and outcomes evaluated. The protocol, materials, statistical analysis scripts and raw data are publicly accessible in our OSF repository (*link*).

RCTs met the following criteria to be eligible: CVR trials containing at least one arm including any type of EBI; (b) and randomized patients with CVD-related morbidities, as follows: coronary artery disease, stable angina, unstable angina, acute myocardial infarction (STEMI and non-STEMI), heart failure independent of the ejection fraction status, primary/secondary hypertension, cerebrovascular disease (ischemic or hemorrhagic stroke), and peripheral arterial disease. In our study, EBI were the ones including aerobic or resistance exercises, supervised (by a professional) or not, individually or in groups, delivered at home (home-based) or in another setting, in patients with diagnosed CVD [15]. Studies with arms in which primary intervention was motivational, nutritional, counseling, or usual care (ie, standard medical care such as medication use or a non-pharmacological, non-structured intervention) were not eligible.

### Search strategy and screening

Our unit of analysis was systematically collected after a previous survey of journals, in which was applied the search strategy to therefore retrieve the eligible manuscripts for the analysis. We queried Scopus database using the “Journals” filter, as well as the fields of interest: “*Physical Therapy, Sports Therapy and Rehabilitation”, “Cardiology and Cardiovascular Medicine”*, and *“Rehabilitation”*. The top 100 journals ranked by Scopus CiteScore 2019 were screened by the following eligibility criteria: (a) scope of interest (i.e., Cardiology and Rehabilitation) and (b) have at least one CVR trial published between 2017 and 2021. We then searched for RCTs of CVR in each eligible journal applying a search strategy in PubMed/MEDLINE, composed by free terms, MeSH terms, and relevant descriptors related to exercise, plus an ultrasensitive filter for RCTs [25] (please see appendix A). 10% of the retrieved articles were piloted to test the eligibility process flow, as well as library management. After the approval of a senior researcher over the pilot sampling, the sample was organized in Zotero 5.0.96 reference manager software.

### Studies Selection

The sample of articles was manually deduplicated with Zotero and alphabetically organized by title. The screening of the eligible studies was conducted in duplicity, with independent and blinded reviewers (GWF + LL and ASK + RZ) tracking the titles and abstracts in the first hand and excluding the ones who had judged as ineligible to a separate folder, named by priority, as follows: 1-“Not humans”; 2 - “Not a RCT”; 3 - “Not in English”; and 4 - “Others” (those with additional ineligible characteristics). The potentially eligible articles were selected after judgement considering the full text. Disagreements were adjudicated by a third investigator. The eligibility process of journals and articles can be viewed in Figure 1.

**Figure 1.**
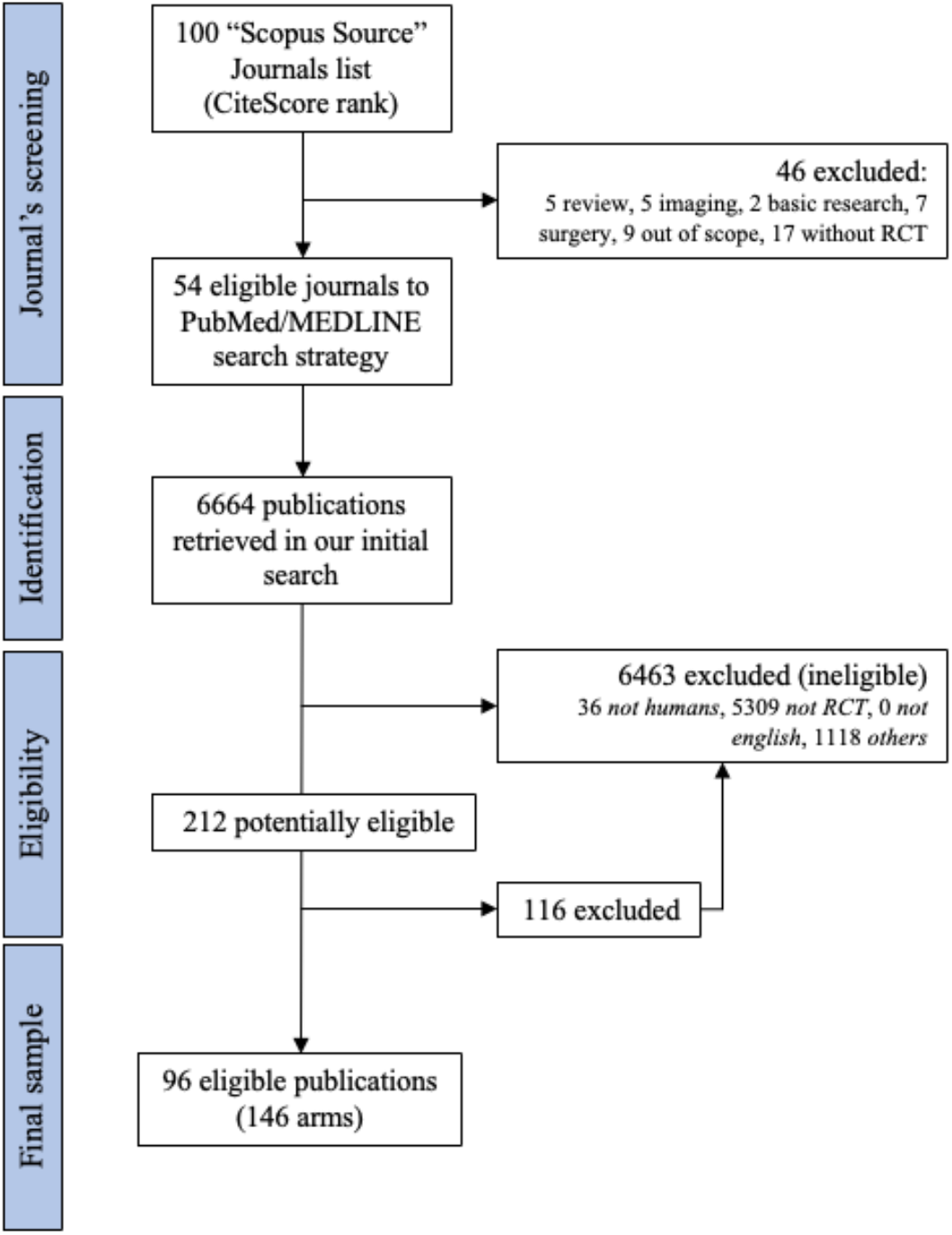
Flowchart of the journals and studies included in the analysis.

### Data extraction

The data collection was carried by the same set of researchers of the screening phase, using a Google form previously created. In order to conduct the extraction training, a single author (GWF) piloted an article [26] via virtual meeting (record available in our public repository), where an explanation of the project’s objectives, methods (structure of the form), training and outcomes was provided. The following studies’ general characteristics were collected: (a) year of publication; (b) impact factor (SJR); (c) journal location (continent); (e) material sharing (e.g., research instruments); (f) data sharing statements (link or URL); (g) raw data sharing statements (code or syntax); (h) link to an available study protocol; (i) preregistration report (e.g., registry number); (j) statement of whether it was a reproducibility study or not; (k) conflict of interest statement; (l) funding statement; (m) open access article.

The prevalence of methods and outcomes reported was assessed with a total of 12 items, which was answered dichotomously with YES or NO, respectively to the presence or absence of reporting (i.e., description or statement) within the eligible primary study; or by counting prevalence (frequency of an item). We collected the following methodological and outcome items: (1) primary outcome effect size description (YES/NO); (2) primary outcome aggregation method description (YES/NO); (3) p-value precisely reported? (YES/NO); (4) statistically significant primary outcome reported? (YES/NO); (5) any standardized effect size (SES) measurement reported (YES/NO); word “significant” prevalence in a statistical context (count); (7) statement of a clinically meaningful result by means of statistical significance (YES/NO); (8) Spin in Title OR Abstract sections (count); (9) Spin in Results section (count); (10) Spin in Conclusion section (count); (11) reporting of statistical power (YES/NO); (12) sample size calculation reported? (YES/NO). We extracted the prevalence of spin in accordance to classification of Boutron et al. [27] in both positive and negative articles (i.e., primary outcome < or > 0.05), considering that they are included in the “other types” category. Additional variables were manually collected: the sample size (i.e., subjects enrolled to randomization); estimated sample size; and whether the primary outcome confidence interval was reported (descripted in the statistical analysis of the results section). These variables were collected after the first version of the protocol, since we understood that they had statistical relationships in potential.

### Statistical analysis

Descriptive statistics for the primary outcome were described as counts and prevalence for categorical variables, within a 95% precision of estimates (95% confidence interval). For continuous variables, we described estimates through means and standard deviations (SD), or medians and, depending on its density distribution.

To answer our main objective, the reporting of both methodological and outcome characteristics was analyzed from the sample of CVR trials. Our primary outcome was the prevalence of these characteristics by means of descriptive statistics for both categorical (frequencies and proportions) and continuous (mean, SD, interquartile intervals, minimum and maximum) variables. For our secondary outcome, we estimated the prevalence ratio (PR) by means of a negative binomial regression model [29], as the response variable (Spin) presented previous overdispersion (variance greater than the mean) [30].

## Results

Primary studies’ general characteristics, as well as the variables related to transparency and reproducibility are shown in Table 1.

**Table 1.** General characteristics of studies and the reporting transparency and reproducibility variables.

| Characteristics | Variables |  |  |
| --- | --- | --- | --- |
| N=96 | Ref. | n (%) | CI 95% |
| Year of publication | 2017 | 26 (27.08) | 19.1 - 36.9 |
|  | 2018 | 18 (18.75) | 12.1 - 27.9 |
|  | 2019 | 22 (22.92) | 15.5 - 32.5 |
|  | 2020 | 27 (28.12) | 19.9 - 38.0 |
|  | 2021 | 3 (3.12) | 1.0 - 9.3 |
| Journal impact factor (SJR) | Mean | 2.35 | 1.9 - 2.5 |
|  | SD | 1.89 |  |
|  | Min-Max | 0.79 - 10.31 |  |
| Journal location | Americas | 41 (42.71) | 45.1 - 64.9 |
|  | Europa | 53 (55.21) | 33.1 - 52.9 |
|  | Oceania | 2 (2.08) | 0.5 - 8.1 |
| It is stated in the article if materials are available? | No | 71 (73.96) | 64.1 - 81.8 |
|  | Yes | 25 (26.04) | 18.1 - 35.8 |
| It is stated in the article if the data are available? | No | 61 (63.54) | 53.3 - 72.6 |
|  | Yes | 35 (36.46) | 27.3 - 46.6 |
| It is stated in the article the analysis scripts are available? | No | 93 (96.88) | 90.6 - 99.0 |
|  | Yes | 3 (3.12) | 1.0 - 9.3 |
| Did the authors shared a link to an accessible protocol? | No | 80 (83.33) | 74.3 - 89.6 |
|  | Yes | 16 (16.67) | 10.4 - 25.6 |
| Is there a statement about whether the article was preregistered or not? | No | 23 (23.96) | 16.4 - 33.6 |
|  | Yes | 73 (76.04) | 66.4 - 83.6 |
| Are there any statements if the article was a replication or a new one? | No | 76 (79.16) | 69.8 - 86.2 |
|  | Yes.new | 20 (20.84) | 13.8 - 30.2 |
|  | Yes.replication | 0 (0.00) |  |
| Does the article include any statements of a conflict of interest? | No | 21 (21.88) | 14.6 - 31.4 |
|  | Yes | 75 (78.12) | 68.6 - 85.4 |
| Does the article include any statements of funding source? | No | 12 (12.50) | 7.2 - 20.8 |
|  | Yes | 84 (87.50) | 79.1 - 92.9 |
| It is an open access article? | No | 37 (38.54) | 29.3 - 48.7 |
|  | Yes | 59 (61.46) | 51.3 - 70.7 |

SD: standard deviation; CI: confidence interval; IQR: interquartile range.

Regarding the reporting of methods and outcomes (table 2), the primary outcome effect size was not reported in 35% of the studies, and 74 (77%) of the 96 studies do not report any SES. However, more than 2/3 of the sample (69%) had a statement in the discussion or conclusion sections mentioning clinical relevance or meaningful benefit of the statistically significant results.

**Table 2.**
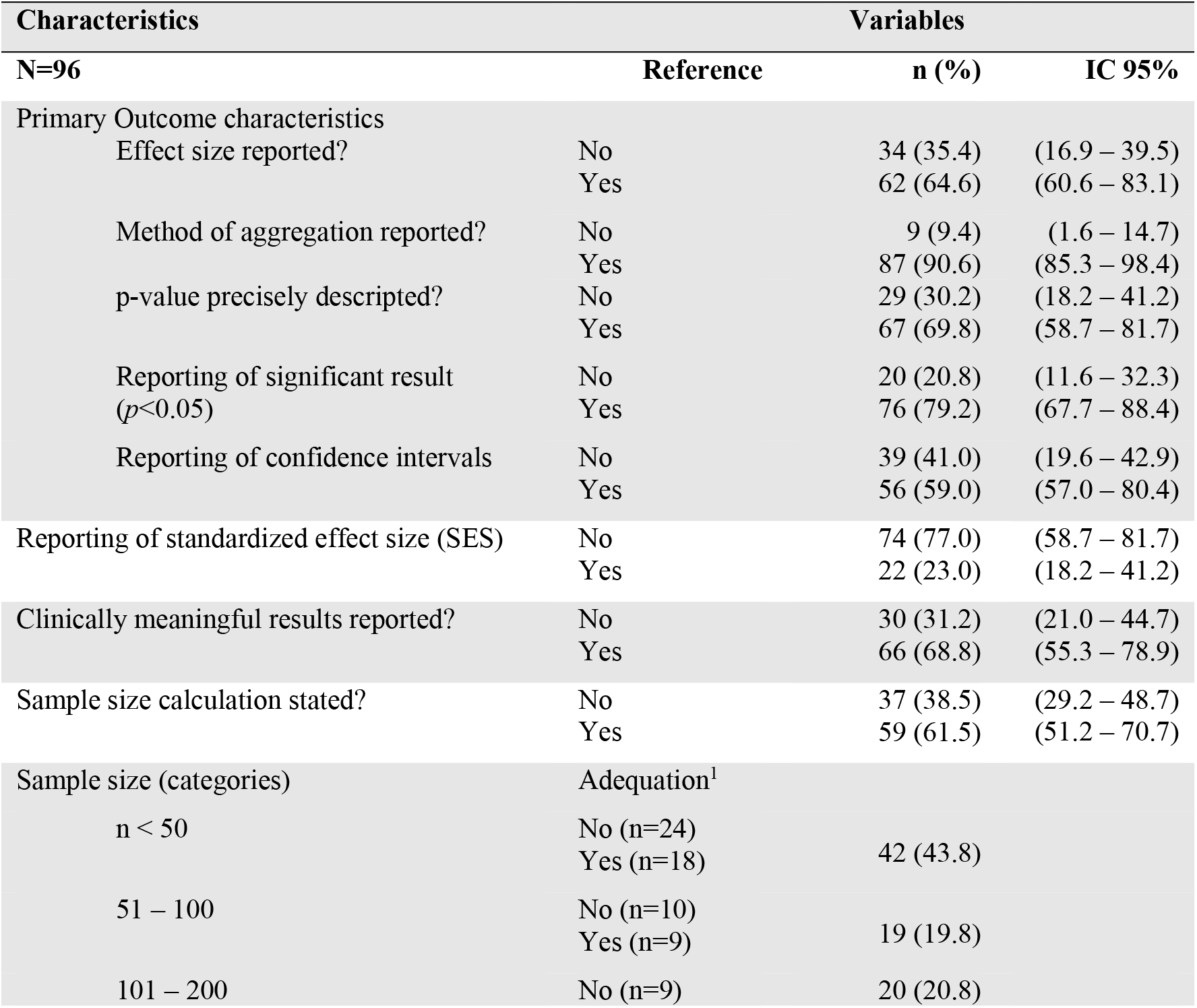
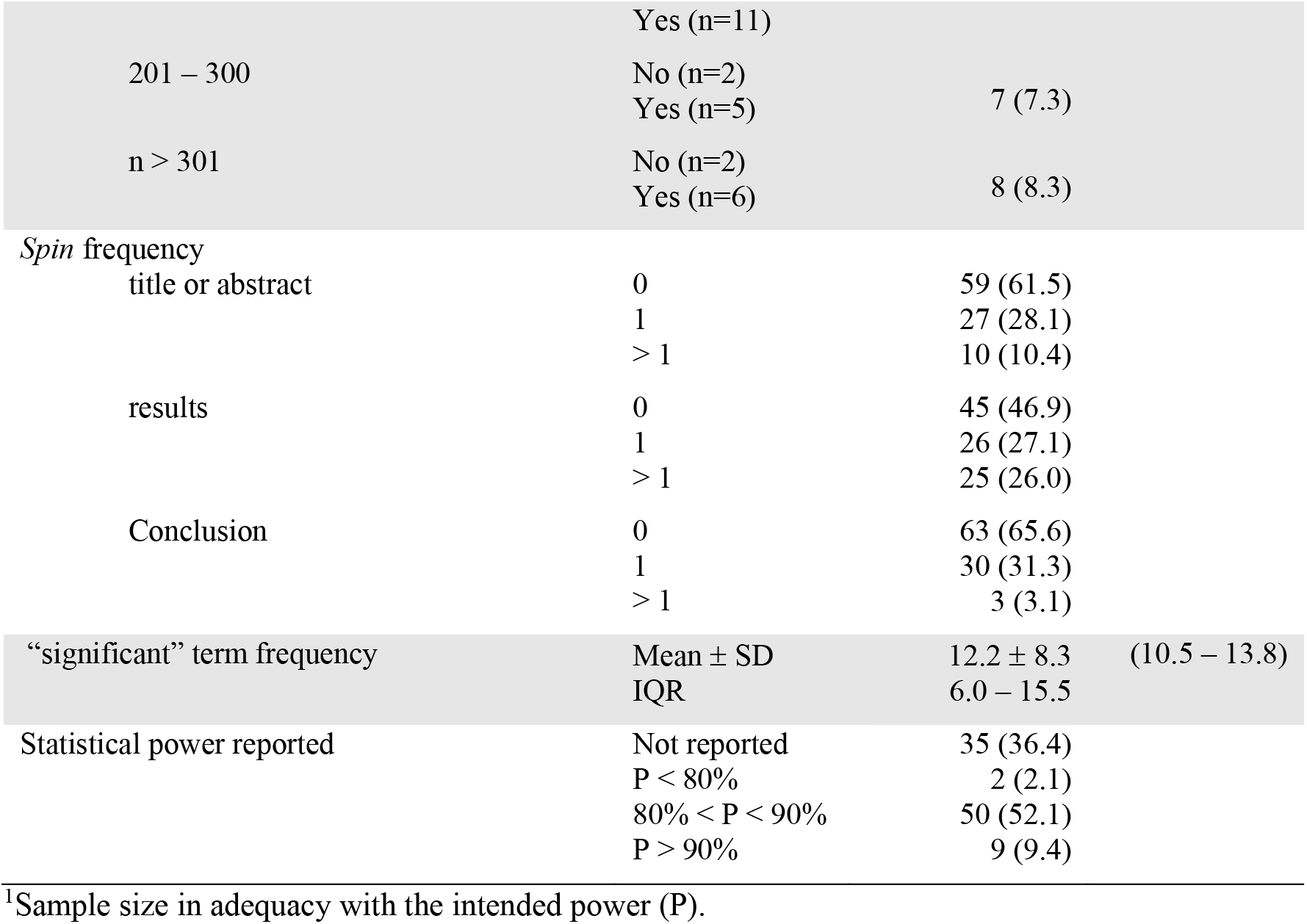
Characteristics of methods and results (i.e., outcomes) reported in studies.

Approximately 60% of the total sample of 96 RCTs had a clear reporting of the sample size calculation, considering the description of the study population, power, significance level, minimum detectable difference, variance, or dropout rate [31]. Half of the sample (51%) showed adequacy to the sample size calculation (i.e., the sample of subjects randomized in the studies was greater than or equal to the calculated sample size), while the other half (49%) either showed inadequacy or did not describe the sample size calculation. In view of the sample size heterogeneity, we categorized the studies. Samples of less than 100 subjects (63%) showed greater inadequacy in relation to sample size calculation, with a major difference in studies with less than 50 randomized subjects (43%), of which 57% do not have a sample consistent with the intended power.

The prevalence of at least one Spin was 67 (70%) of the 96 studies, considering the title or abstract sections, results, and conclusions together. The results section showed a higher frequency, of which 51 (53%) had at least one Spin. The conclusion section had a lower prevalence of Spin, in which 63 (66%) of the articles did not report any type of Boutron’s spin types. The term “significant” and its derivations (e.g., “significantly”, “near significant”, “quasi-significant”) appeared in the results and discussion/conclusion sections with a median of 11 (in a statistical outcome context).

Our secondary outcome sought relationships between the reporting of any measure of effect size with the amount of Spin, considering the studies that did not declare the primary outcome as statistically significant. An association was found between the variables, as shown in the upper portion of the table 3. Thus, the reporting of effect size may interfere in the prevalence of Spin reported in the results section (PR=0.11; *P*=0.01; 95%CI 0.03-0.44) and total of spin (PR=0.34; *P*=0.01; 95%CI 0.15-0.77), among the studies that reported a non-significant primary outcome. In contrast, reporting a primary outcome as statistically significant was not associated with the prevalence of Spin when compared with negative studies. In other words, the direction of the results (i.e., authors declaring the primary outcomes as significant or not) does not seem to be explained by the prevalence of Spin.

**Table 3.** Association between the effect size reporting and the primary outcome direction with the prevalence of spin.

| Outcome variable | Comparison | PR* | CI 95% | P |
| --- | --- | --- | --- | --- |
| Spin prevalence | Reporting Prevalence | PR* | CI 95% | P |
| **Studies with PO p>0.05** |  |  |  |  |
| Spin in title or abstract | Effect size (Yes vs. No) | 0.57 | 0.19-1.70 | 0.31 |
| Spin in results | Effect size (Yes vs. No) | 0.11 | 0.03-0.44 | 0.01 |
| Spin in conclusion | Effect size (Yes vs. No) | 1.55 | 0.40-6.01 | 0.52 |
| Total of spin | Effect size (Yes vs. No) | 0.34 | 0.15-0.77 | 0.01 |
| **Studies with PO p<0.05** |  |  |  |  |
| Spin in title or abstract | Effect size (Yes vs. No) | 1.13 | 0.53-2.42 | 0.74 |
| Spin in results | Effect size (Yes vs. No) | 0.97 | 0.52-1.79 | 0.92 |
| Spin in conclusion | Effect size (Yes vs. No) | 0.98 | 0.43-2.20 | 0.96 |
| Total of spin | Effect size (Yes vs. No) | 1.00 | 0.58-1.74 | 0.97 |
| Spin in title or abstract | PO p<0.05 (Yes vs. No) | 0.71 | 0.38-1.39 | 0.29 |
| Spin in results | PO p < 0.05 (Yes vs. No) | 0.84 | 0.42-1.64 | 0.61 |
| Spin in conclusion | PO p < 0.05 (Yes vs. No) | 0.68 | 0.34-1.49 | 0.31 |
| Total of spin | PO p < 0.05 (Yes vs. No) | 0.77 | 0.44-1.32 | 0.35 |

\* Negative binomial regression model; PO: primary outcome; PR: prevalence ratio; CI: confidence interval

## Discussion

Our study shows that CVR trials with EBI insufficiently reported methods and outcomes. Studies reporting effect size measures had a lower chance of Spin. More than half of the sample presented at least one spin in the results section, which is a similar prevalence in comparison to what is found in the biomedical literature [28], as well as in the cardiovascular field [24].

Inadequate statistical reporting may misguide the interpretation of the results and is also prevalent in the CVR field. Some of the most concerning are the lack of statistical power, the misuse of statistical analysis, and absence of explanation about atypical or missing data [21]. Furthermore, it is difficult to differentiate outcomes with small effect sizes from biases, which turns the effect questionable even when the result is statistically significant [32]. Researchers should report the effect sizes to facilitate the interpretation of a SRMA (e.g., when comparing standardized effect-sizes across studies), as well as the sample power calculations on which independent authors rely to develop new studies [33]. Recently, Chu et al. [34] analyzed the statistical reporting from a sample of 6124 studies published in highly ranked biomedical journals within the last decade. They showed that only 24% of the studies properly reported the effect size. As far as we know, our study is one of the few describing the prevalence of effect size reporting within a sample of CVR trials, in which 35% of studies failed in this regard.

Confidence intervals (CI) should be clearly reported by the researchers as well, as it provides essential information about the inferential probability and conclusions regarding the clinical relevance of the findings, besides being a better estimate of the effect than the reported p-values itself [35]. Although nearly 70% studies had stated its outcomes as clinically meaningful within our sample, about 41% (39/96) of the studies clearly stated the primary outcome CI. Even so, it is necessary to consider the importance of the accurate description of the p-value [18], found in our sample with a prevalence of 80%. However, it should be noted that statistical significance can be determined by assessing whether the defined limits (e.g., alpha level < 0.05) are within the CI range [36]. A clinically meaningful result is an estimative that exceeds a minimally important threshold for the patient to perceive the benefit in order to deliver a evidence-based clinical decision [37]. The reporting of inferential statistical aspects in the CVR field have been briefly studied [38], showing a prevalence ranging from 15% up to 86. It is recommended to report the CI accompanied by its effect sizes rather than reporting the p-value alone, which can be a feasible strategy for preventing Spin [39].

Spin are practices in which the author states redundant description of results trying to explain unfavorable or favorable findings, or even emphasizing favorable results, in addition to strategies for statistical presentation of results without prior rationale [28]. These practices are more prevalent in RCTs, especially when the superiority null hypothesis is rejected (i.e., non-significant primary outcome) [28]. In our sample, the prevalence of spin in the results section is higher among those trials that reported the effect size (PR=0.12; P<0.01; 95%CI 0.03-0.41). However, we did not found association regarding the direction of the primary outcome and the prevalence of spin, contrary to what have been found in the literature [27,40–43]. Nevertheless, in a SRMA evaluating the level and prevalence of spin in the CV literature (KHAN et al., 2019), the authors found that 67% of RCTs presented a misuse of language resources among studies with non-significant results and driving the arguments mainly to significant secondary outcomes. Within our sample, we found a similar prevalence of 70% (table 2). This could be explained by the wide variation in the impact factor within our sample, which includes journals in which the editorial and peer-review policies may not be as stringent as those highly ranked. Still, those authors found an association between the spin level in the abstracts and the primary outcome but no association between the spin level in the body of the article and the primary outcome, contrary to what was found in our analysis (please see the bottom lines in table 3).

There is a worrying prevalence of sample size calculation reporting in the rehabilitation literature [44], and this is important because underpowered studies are unable to detect relevant clinical effects. A survey with 215 RCTs published in high-impact medical journals [7], showed that only 1/3 adequately describe (i.e., sufficiently to allow replication) the sample size calculations. Within the CVR pre-clinical studies with therapeutic intervention, only 4.2% properly reported the sample power calculations [45]. Regarding our sample, 40% (37/96) of the studies did not report *a priori* sample size calculation. Furthermore, 1/4 (34%) of the studies did not state the number of randomized subjects that could meet the intended power. To our knowledge, this is the first study assessing these characteristics altogether in the exercise based CVR literature.

Regarding the limitations of this study, we extracted Spin prevalence according to its report in the different sections of the article without a verification of the most prevalent spin strategies and its types [27]. The prevalence of spin from both positive and negative studies was extracted, including those studies where a redundant description of results led to an explanation of unfavorable findings or those where favorable outcomes was highlighted. Another limitation that may have biased our results is that we only relied on the training meeting to seek extraction agreement, not running a Kappa statistic for example. In this study, we descriptively explored the reporting of variables and did not incur in meta-regression (e.g., associations between magnitudes of effects), which does not add inferential evidence to the literature. Outcome reporting items (e.g., spin) can be very subjective to interpretation, which may have biased our analysis. We were also unable to assess the effect of spin on reviewers, editors, and health professionals, since it comprised a limited period in the selected journals, which does not allow us to better understand the scenario of the last decade in terms of the quality of reporting outcomes and methodological features of exercise-based CVR trials.

## Conclusion

Our study found a low prevalence of methodological and outcome reporting characteristics in primary studies of exercise-based CVR trials. Selective outcome reporting in RCTs has important implications for translating science into practice, as it threatens the interventions effectiveness validity. Also, these practices frustrate the use of its evidence in meta-analyses, thus hampering the synthesis of evidence that guides clinical practice and the development of future studies and health policies.

## Data Availability

All data produced are available online indefinitely at https://osf.io/754ht/

## Appendix

### A. Search strategy

Randomized Controlled Trial Filter [25]

(randomized controlled trial [pt] OR controlled clinical trial [pt] OR randomized controlled trials [mh] OR random allocation [mh] OR double-blind method [mh] OR single-blind method [mh] OR clinical trial [pt] OR clinical trials [mh] OR (“clinical trial” [tw]) OR ((singl* [tw] OR doubl* [tw] OR trebl* [tw] OR tripl* [tw]) AND (mask* [tw] OR blind* [tw])) OR (“latin square” [tw]) OR placebos [mh] OR placebo* [tw] OR random* [tw] OR research design [mh:noexp] OR comparative study [mh] OR evaluation studies [mh] OR follow-up studies [mh] OR prospective studies [mh] OR cross-over studies [mh] OR control* [tw] OR prospectiv* [tw] OR volunteer* [tw]) NOT (animal [mh] NOT human [mh]))

PubMed/MEDLINE

#1 (“Title of the journal”[Journal])

#2 (Cardiac Rehabilitation[Mesh] OR Exercise Therapy[Mesh] OR Sports[Mesh] OR Physical Exertion[Mesh] OR rehabilitation OR (physical* AND (fitness or training or therapy or activity)) OR Exercise[Mesh] OR (train*[tiab](strength[tiab] or aerobic[tiab] or exercise[tiab])) OR (exercise[tiab] or fitness[tiab]))

#3 (treatment[tiab] or intervention[tiab] or program[tiab])) OR Rehabilitation[Mesh] OR kinesiotherapy

#1 AND #2 AND #3

## Additional Information

### Compliance with Reproducibility and Data Sharing Standards

This project is in accordance with the current standards of transparency indicated by the International Committee of Journal Medical Editors, the Committee of Publication Ethics, has a Principal Investigator (PI) as a signatory of the San Francisco Declaration of Research Assessment (DORA) and endorses the Hong Kong Manifesto for the assessment of researchers, faculty, and others. This project intends to publish all its scientific pieces in a pre-print version and an open-access journal, whenever possible.

Independent authors will have full access to our data including: Zotero libraries with eligible and ineligible articles, statistical codes used in analysis, statistical analysis, glossary of variables and protocol in our public repository without time constraints nor request conditions. The data will be available immediately following publication with no end date. Data is available indefinitely (https://osf.io/754ht/).

### Funding Sources

This project has not received funding from any source to be conducted.

### Author’s contributions

Conception: GWF

Data Curation: GWF

Formal Analysis: GWF

Investigation: GWF, RZ, LL, ASK

Methodology: GWF, TC

Data treatment: GWF

Writing: GWF

Revision: TC, RZ, ASK

Final edition: GWF

Supervision: TC

Project administration: GWF

Funding acquisition: N/A.

## Notes

### Competing Interest Statement

The authors have declared no competing interest.

### Funding Statement

This study did not receive any funding

